# Deciphering TB-IRIS in HIV Patients: A Comprehensive Clinical and Microbiological Analysis

**DOI:** 10.1101/2024.09.10.24313396

**Authors:** Priyanka Gupta, Anil Kumar Tripathi, Kaleshwar Prasad Singh, Amita Jain, Abhishek Gupta

## Abstract

**Background:** Tuberculosis (TB) and Human Immunodeficiency Virus (HIV) coinfection presents a complex clinical challenge, with Tuberculosis-Immune Reconstitution Inflammatory Syndrome (TB-IRIS) emerging as a significant complication following antiretroviral therapy (ART) initiation. Understanding the clinical and microbiological characteristics of TB-IRIS is crucial for improving patient outcomes.

**Methods:** This prospective cohort study included 400 HIV-positive patients from the ART centre at King George’s Medical University, Lucknow, India. Patients were categorized based on TB status and monitored for the development of TB-IRIS following ART initiation. Clinical data, CD4 counts, and microbiological analyses, including drug susceptibility testing, were conducted. TB-IRIS was classified as paradoxical or unmasking, and outcomes were assessed over a one-year period.

**Results:** Among the 400 patients, 38 (9.5%) developed TB-IRIS, with 31 (81.6%) presenting unmasking TB-IRIS and 7 (18.4%) paradoxical TB-IRIS. Tubercular meningitis (TBM) was the most common manifestation (47.3%), followed by pulmonary TB (29.0%). The incidence of TB-IRIS was higher (15.4%) in patients who initiated ART within one month of starting anti-tuberculosis therapy (ATT) compared to those who started ART later (5.5%). A lower baseline CD4 count (<100 cells/µL) was significantly associated with a higher risk of TB-IRIS (p=0.003). The drug resistance analysis revealed 27.2% resistance to both isoniazid and rifampicin. Steroid therapy was administered to 13% of TB-IRIS patients. The overall cure/improvement rate was 71%, while the mortality rate was 23.6%. No patients discontinued ART during the study.

**Conclusion:** This study highlights the predominance of unmasking TB-IRIS in HIV patients initiating ART, particularly those with low baseline CD4 counts and early ART initiation post-ATT. The significant drug resistance observed underscores the need for robust diagnostic and treatment protocols. Improved management strategies are essential to enhance clinical outcomes in TB-IRIS patients.

## Introduction

The care and treatment of patients might be complicated by the intersection of two significant global health concerns, namely tuberculosis (TB) and HIV. In addition to being the most common cause of morbidity and death among those with HIV, TB was the main cause of mortality. Conversely, TB can accelerate the progression of HIV, creating a complex clinical scenario that requires nuanced management strategies. [1]. One of the significant complications arising from the co-infection of HIV and TB is TB-Immune Reconstitution Inflammatory Syndrome (TB-IRIS). TB-Immune Reconstitution Inflammatory Syndrome, is a condition that can occur in people who have HIV and start antiretroviral therapy (ART) [2,3]. It happens when the immune system begins to recover from HIV infection and, as a result, mounts an inflammatory response against a previously treated or latent TB infection.

TB-IRIS is classified into several types based on when the symptoms appear and their severity; Paradoxical TB-IRIS: This type occurs when the symptoms of TB worsen after starting ART, despite proper TB treatment. The immune system’s recovery can lead to an exaggerated inflammatory response to TB that was previously under control [4,5]. Unmasking TB-IRIS: This type occurs when new TB symptoms or signs emerge after starting ART, which were previously asymptomatic or not detected. The immune system’s recovery can reveal previously hidden or subclinical TB infection. [2,6]. The management of TB-IRIS typically involves continuing both TB treatment and ART, along with symptomatic management and sometimes corticosteroids to control inflammation. Regular monitoring and clinical evaluation are crucial to ensure appropriate care.

The clinical management of TB-IRIS presents a formidable challenge due to its unpredictable nature and the overlap of symptoms with both TB and ART-related side effects. Understanding the clinical manifestations and microbiological underpinnings of TB-IRIS is crucial for developing effective treatment strategies and improving patient outcomes [7].

This study aims to characterize TB-IRIS in HIV patients by exploring both clinical and microbiological dimensions. By examining the specific clinical features associated with TB-IRIS and the interactions between *Mycobacterium tuberculosis (M*.*tb*.) and the recovering immune system, this research seeks to enhance the understanding of this complex syndrome. The findings are expected to contribute to better diagnostic criteria, treatment protocols, and ultimately, improved management of co-infected patients. Through this research, we hope to address existing knowledge gaps and provide insights that will benefit clinicians in managing TB-IRIS, offering a more refined approach to treating patients who face the dual burden of HIV and TB.

## Materials and Methods

### Study population

In a prospective cohort study, 400 HIV positive patients were included. The study subjects were recruited from ART centre, King George Medical University (KGMU), Lucknow, India (Figure 1). The inclusion criteria used for the selection of the subjects were: 1) Patients diagnosed having HIV infection, 2) ART eligibility according to National AIDS Control Organization (NACO) Guidelines, 3) ART naive, 4) HIV positive patients already on ATT and 5) Willingness to participate in the study and exclusion criteria: 1) Patient not willing to give consent, and 2) Patients who were already on ART. The study experiment and data accumulation were carried out with the understanding and informed consent of the human subject and with approval from Institutional ethical committee, KGMU, Lucknow, India.

**Figure 1.**
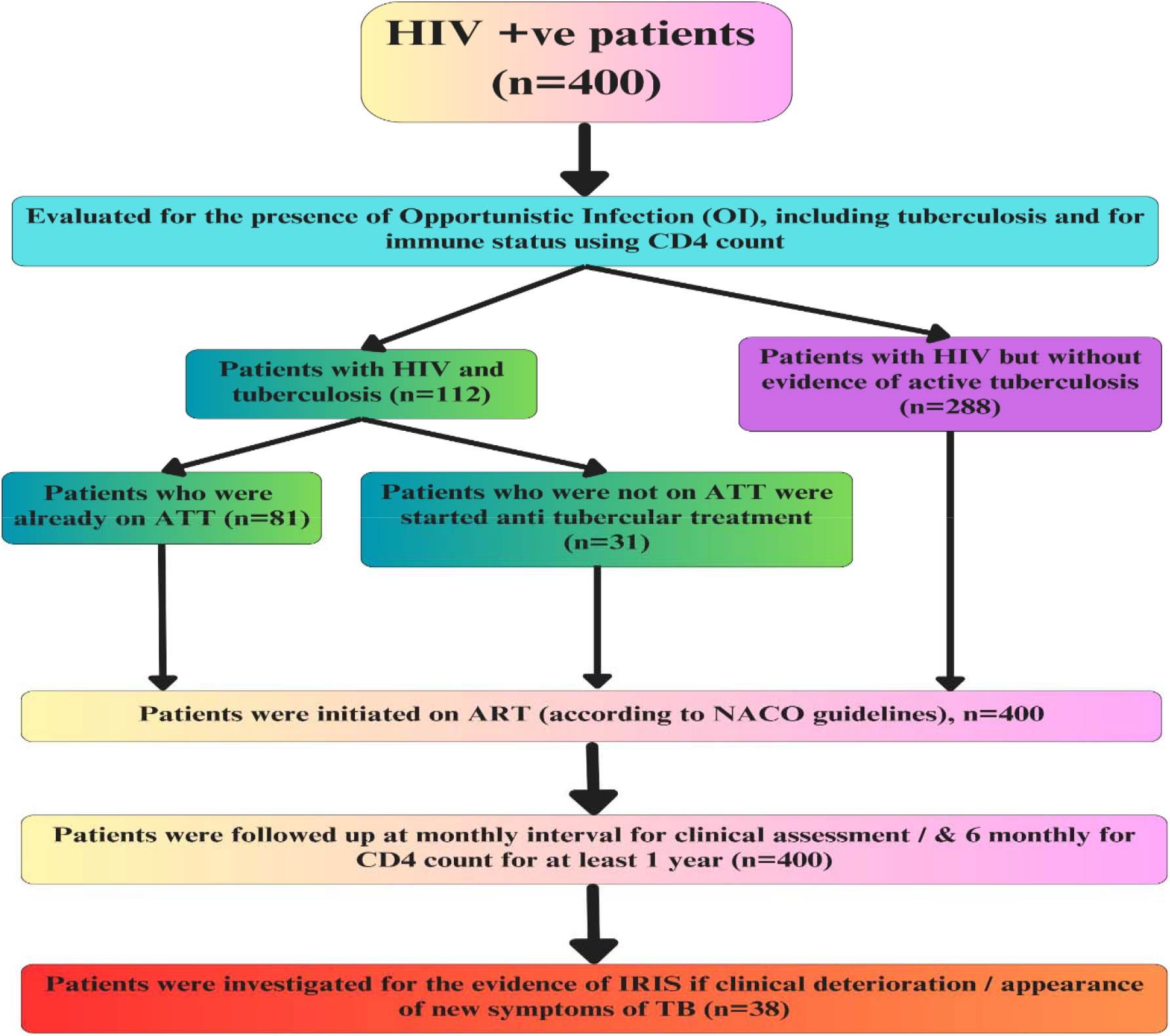
Flow chart of TB-IRIS in HIV +ve patients.

### Clinical data measurements

All patients were diagnosed as HIV positive using ELISA according to NACO guidelines (2011) [8]. A pre-designed structured Performa was used to collect socio-demographic characteristics and relevant clinical data of the patients, and their information related to risk factors for acquiring IRIS. All patients subjected to thorough clinical history and examinations.

### Diagnosis of active TB

Patients were considered to have active TB when they had a two positive sputum smears by AFB, positive mycobacterium culture on Lowenstein Jensen media and if they had TB symptoms (any one of cough of more than 2 weeks, fever, loss of appetite, night sweats or loss of weight). Patients were divided into two groups: 1) HIV patients with active TB who were receiving ATT, and 2) HIV patients with no evidence of active TB. Tuberculin skin test was also performed to assess *in vivo* immune response to TB. Culture and drug susceptibility testing (DST) for *M*.*tb*. were done in all cases where specimens were available to rule out drug-resistant TB.

### CD_4_ cell count

We collected blood for CD_4_ cell count measurement at enrolment and at month six follow up visit. Lymphocyte subsets (CD_4_ cell counts) were analysed by standard three-color flow cytometry (FACScan; Becton Dickinson, San Jose, CA, USA).

### Statistical analysis

Continuous variables such as age, Hb and CD_4_ cell count were summarized into means and medians as appropriate with t test, paired and unpaired as appropriate. The 95% confidence intervals were constructed around the estimates and the p-values used as a measure of statistical significance. A p-value of 0.05 or less was considered significant. Data analysis was done using SPSS 15.0 version statistical software.

## Results

The majority of TB-IRIS cases were new (“unmasking”) presentations, comprising 81.6% (31 patients). Worsening (“paradoxical”) TB-IRIS was observed in 18.4% (7 patients). Tubercular Meningitis (TBM) was the most common presentation, affecting 47.3% (18 patients). Pulmonary TB was observed in 29.0% (11 patients). Cervical Lymphadenopathy was seen in 8.0% (3 patients). Abdominal Lymphadenopathy affected 13.0% (5 patients). Disseminated TB was the least common presentation, found in 2.7% (1 patient). Only 13% (5 patients) received steroid therapy for TB-IRIS. The majority, 87% (33 patients), did not receive steroid therapy. None of the patients (0%) discontinued antiretroviral therapy (ART) during the study period. A positive outcome, defined as cured or improved, was achieved in 71% (27 patients) of cases. There were 9 deaths (23.6%) among the patients. A small proportion of patients, 2.63% (1 patient), were lost to follow up (LFU). Another 2.63% (1 patient) were transferred out during the study (Table 1).

**Table 1.**
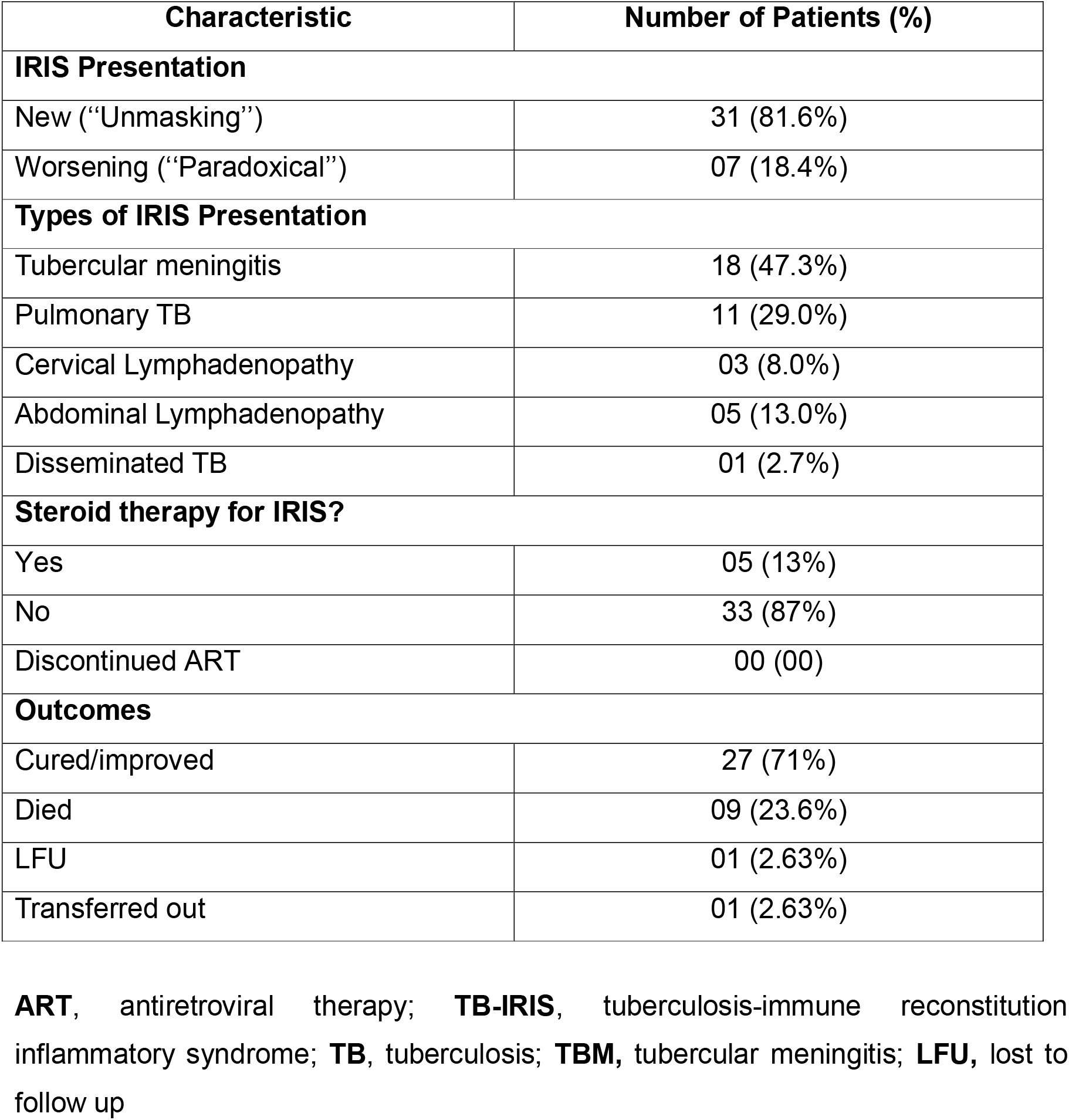
Clinical manifestations, treatment, and outcomes of TB-IRIS cases.

| Characteristic | Number of Patients (%) |
| --- | --- |
| **IRIS Presentation** |  |
| New (“Unmasking”) | 31 (81.6%) |
| Worsening (“Paradoxical”) | 07 (18.4%) |
| **Types of IRIS Presentation** |  |
| Tubercular meningitis | 18 (47.3%) |
| Pulmonary TB | 11 (29.0%) |
| Cervical Lymphadenopathy | 03 (8.0%) |
| Abdominal Lymphadenopathy | 05 (13.0%) |
| Disseminated TB | 01 (2.7%) |
| **Steroid therapy for IRIS?** |  |
| Yes | 05 (13%) |
| No | 33 (87%) |
| Discontinued ART | 00 (00) |
| **Outcomes** |  |
| Cured/improved | 27 (71%) |
| Died | 09 (23.6%) |
| LFU | 01 (2.63%) |
| Transferred out | 01 (2.63%) |

**ART**, antiretroviral therapy; **TB-IRIS**, tuberculosis-immune reconstitution inflammatory syndrome; **TB**, tuberculosis; **TBM**, tubercular meningitis; **LFU**, lost to follow up

### Prevalence of IRIS in relation to TB status

Out of 81 patients diagnosed with active TB, 7 (8.6%) were diagnosed with IRIS. Among the 319 patients without TB, 31 (9.7%) were diagnosed with IRIS. The relative risk (RR) of developing IRIS for patients with active TB compared to those without TB was calculated to be 0.89 (95% Confidence Interval [CI]: 0.41– 1.95), with a p-value of 0.77 (Table 2). This indicates that the risk of IRIS in patients with active TB is not significantly different from the risk in those without TB.

**Table 2.** Prevalence of IRIS in relation to TB status.

| Variables | Number of patients (n=400) | Prevalence of IRIS |  | RR (95% CI)<br>p value |
| --- | --- | --- | --- | --- |
|  |  | No. | % |  |
| Active TB | 81 | 7 | 8.6 | 0.89 (0.41-1.95)<br>0.77 |
| Without TB | 319 | 31 | 9.7 |  |

**TB**, tuberculosis; **IRIS**, immune reconstitution inflammatory syndrome; **RR**, relative risk; **CI**, confidence interval

### Timing of starting ART in relation to ATT and the development of IRIS

Fifty-five patients were having ATT for more than 1 month before initiation of ART and 26 patients were initiated on ATT within 1 month of ART. It was found that incidence of IRIS was more (15.4%) in patients where ART was started <1 month of starting ATT. There was no difference in the incidence of paradoxical IRIS if ART was started after one month of ATT (Table 3).

**Table 3.** Timing of starting ART in relation to ATT and the development of IRIS.

|  | No. of patients (n=81) | IRIS developed (n=7) |  |
| --- | --- | --- | --- |
|  |  | No. | % |
| < 1 month | 26 | 4 | 15.4 |
| >=1 month | 55 | 3 | 5.5 |

p=0.13 (non-significant)
**ART**, antiretroviral therapy; **ATT**, anti-TB therapy; **TB**, tuberculosis; **IRIS**, immune reconstitution inflammatory syndrome; **RR**, relative risk; **CI**, confidence interval

### CD4 count at baseline and development of IRIS

The development of IRIS was significantly higher (p=0.003) among those patients whose CD4 count at baseline was <100 (16.8%) as compared to 100-199.9 (9.3%) and >=200 (4.2%) (Table 4).

**Table 4.** CD4 count at baseline and development of IRIS.

| CD4 count | No. of patients | IRIS Developed |  |
| --- | --- | --- | --- |
|  |  | No. | % |
| $<100$ | 107 | 18 | 16.8 |
| 100-199.99 | 150 | 14 | 9.3 |
| $\geq 200$ | 143 | 6 | 4.2 |

$\chi^2=11.35$ , $p=0.003^*$ (Significant)
**CD4**, cluster differentiation; **IRIS**, immune reconstitution inflammatory syndrome

### Clinical Characteristics of New and Worsening TB-IRIS Cases

The average age of patients with new TB-IRIS was 33.06 ± 6.88 years, while those with worsening TB-IRIS had an average age of 37.00 ± 8.21 years. Patients with new TB-IRIS had a baseline CD4 count of 115.32 ± 69.00 cells/ µL, compared to 90.00 ± 89.50 cells/µL in those with worsening TB-IRIS. At the time of IRIS diagnosis, the CD4 count was 257.68 ± 62.22 cells/µL for new TB-IRIS cases and 265.86 ± 117.91 cells/µL for worsening cases. Hemoglobin levels were 9.67 ± 2.28 g/dL in the new TB-IRIS group and 9.22 ± 2.01 g/dL in the worsening TB-IRIS group. 12 patients with new TB-IRIS were smear-positive compared to 1 patient with worsening TB-IRIS. Nineteen patients with new TB-IRIS were smear-negative compared to 6 in the worsening group. Fifteen patients in the new TB-IRIS group were culture-positive versus 1 in the worsening group. Sixteen new TB-IRIS patients were culture-negative compared to 6 in the worsening group. TBM was observed in 13 new TB-IRIS cases and 5 worsening TB-IRIS cases. Ten new TB-IRIS patients had pulmonary TB compared to 1 in the worsening group. Two patients with new TB-IRIS presented with cervical lymphadenopathy versus 1 in the worsening group. This was observed in 5 new TB-IRIS cases and none in the worsening group. 1 new TB-IRIS patient had disseminated TB, with no cases in the worsening group.

Twenty-three patients with new TB-IRIS were cured compared to 4 with worsening TB-IRIS. There were 6 deaths in the new TB-IRIS group and 3 in the worsening group. 1 new TB-IRIS patient was lost to follow up, with no such cases in the worsening group. 1 patient with new TB-IRIS was transferred out, with none in the worsening group.

**Table 5.** Clinical Characteristics of New and Worsening TB-IRIS Cases.

| Characteristic | New<br>("Unmasked")<br>IRIS (N=31) | Worsening<br>("Paradoxical")<br>IRIS (N=07) | p value |
| --- | --- | --- | --- |
| Age (yrs) | 33.06±6.88 | 37.00±8.21 | 0.001** |
| Baseline CD4 count | 115.32±69.00 | 90.00±89.50 | 0.001** |
| CD4 count at time of IRIS | 257.68±62.22 | 265.86±117.91 | 0.001** |
| Hemoglobin (Hb) | 9.67±2.28 | 9.22±2.01 | 0.001** |
| **Clinical manifestations** |  |  |  |
| Smear-positive, sputum | 12 | 01 |  |
| Smear-negative, sputum | 19 | 06 |  |
| Culture-positive | 15 | 01 |  |
| Culture-negative | 16 | 06 |  |
| **Types of IRIS Presentation** |  |  |  |
| Tubercular meningitis | 13 | 05 |  |
| Pulmonary Tuberculosis | 10 | 01 |  |
| Cervical<br>Lymphadenopathy | 02 | 01 |  |
| Abdominal<br>Lymphadenopathy | 05 | 00 |  |
| Disseminated TB | 01 | 00 |  |
| **Outcomes** |  |  |  |
| Cured | 23 | 04 |  |
| Death | 06 | 03 |  |
| Lost to follow up (LFU) | 01 | 00 |  |
| Transferred out | 01 | 00 |  |

**TB**, tuberculosis; **TB-IRIS**, tuberculosis-immune reconstitution inflammatory syndrome; Hb, hemoglobin; CD4, cluster differentiation 4; TBM, tubercular meningitis; LFU, lost to follow up

### Drug sensitivity result of *M. tb*. strains against first line drugs

This table illustrates the distribution of resistance and sensitivity among the patients for the listed drugs. The streptomycin was resistant in 9%. However, isoniazid and rifampicin were resistant in 27.2% each. Ethambutol was resistant in 11.3% (Table 6).

**Table 6.**
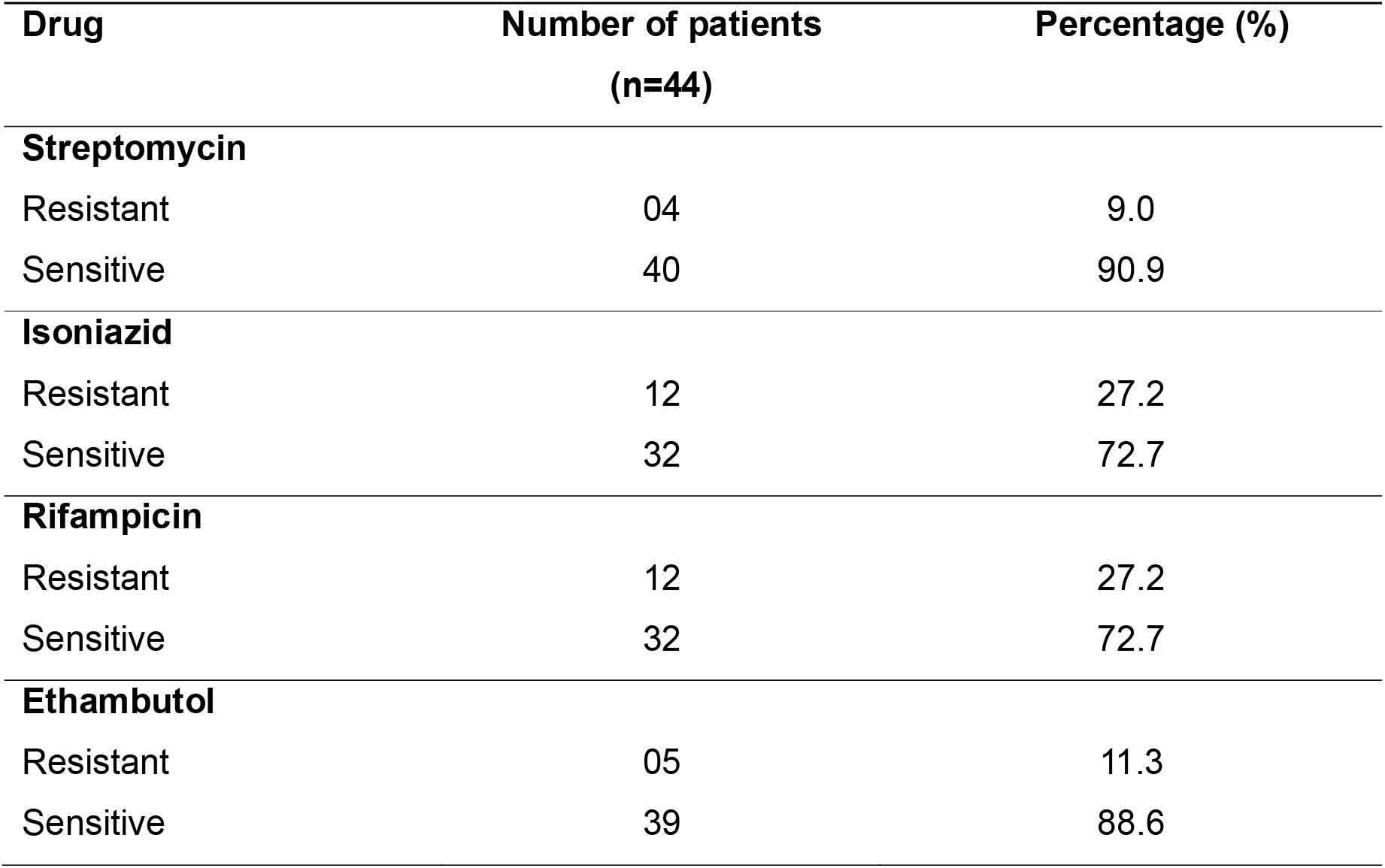
Sensitive and resistant pattern by different drugs.

## Discussion

This study provides a comprehensive look into the characteristics, treatment, and outcomes of patients with TB-Immune Reconstitution Inflammatory Syndrome (TB-IRIS). Our analysis reveals that the majority of TB-IRIS cases were new (“unmasking”) presentations, accounting for 81.6% of cases. This finding, is consistent with earlier studies that have noted a predominance of unmasking TB-IRIS in HIV-infected patients [9,10]. Unmasking TB-IRIS occurs when previously undiagnosed TB becomes clinically apparent following ART initiation. This is contrasted with paradoxical TB-IRIS, observed in 18.4% of patients, where pre-existing TB worsens after ART begins.

The high prevalence of tubercular meningitis (TBM) as the most common presentation (47.3%) aligns with other studies highlighting TBM as a severe manifestation of TB-IRIS [11]. Pulmonary TB, though less common in this cohort (29.0%), is still a significant concern. The relatively low incidence of disseminated TB (2.7%) in our study might reflect advances in early diagnosis and management of TB-IRIS [12].

Steroid therapy for TB-IRIS was administered to only 13% of patients, a notably low percentage compared to historical practices. Historically, corticosteroids were commonly used to manage severe cases of TB-IRIS [13]. The shift away from routine steroid use may reflect updated guidelines and a better understanding of TB-IRIS pathophysiology, which emphasize individualized treatment approaches [14]. The fact that none of the patients discontinued ART during the study underscores the importance of maintaining ART despite IRIS-related complications, as discontinuation can lead to worse outcomes [15].

Our findings suggest that initiating ART within less than one month of starting anti-TB therapy (ATT) is associated with a higher incidence of IRIS (15.4%) compared to those who started ART one month or more after ATT (5.5%). This aligns with previous studies indicating that early ART initiation can increase the risk of IRIS [10, 16-18]. This might be attributed to a more pronounced immune reconstitution effect when ART is started too soon after ATT, leading to heightened inflammatory responses.

The association between lower baseline CD4 counts and higher incidence of IRIS (p=0.003) is consistent with existing literature, which shows that patients with advanced immunosuppression are at higher risk for IRIS [14, 19, 20]. The elevated risk for those with CD4 counts <100 cells/µL highlights the need for careful monitoring and potentially more aggressive management in this high-risk group.

Comparing new and worsening TB-IRIS cases, we observe that new TB-IRIS patients were younger and had higher baseline CD4 counts than those with worsening TB-IRIS. The differences in clinical manifestations and outcomes between these groups emphasize the variability in TB-IRIS presentations and underscore the need for tailored management strategies [9, 11].

The drug resistance patterns observed in our study reveal a significant proportion of resistance to isoniazid and rifampicin (27.2% each), which are critical first-line drugs for TB treatment. This finding underscores the need for routine drug susceptibility testing and highlights the challenges in managing TB-IRIS in the context of drug-resistant TB strains [21, 22]. A great approach to restrict the TB epidemic is to further develop more practical, rapid, and sensitive molecular diagnostic tools to identify subclinical HIV co-infection [23]. HIV co-infected patients can be treated with modern molecular diagnostic tests since they are either accurate or quick enough [24]. HIV patients at risk for TB-IRIS can be predicted using the reliable, sensitive, and easy-to-use tuberculin test (TST) [25].

The high cure or improvement rate (71%) observed in this study is encouraging and aligns with recent findings by [26], who reported favourable outcomes with appropriate management of IRIS. The mortality rate of 23.6% is consistent with some recent studies but is notably high, indicating the need for more effective management strategies [27].

## Conclusion

This comprehensive analysis of TB-IRIS in HIV patients underscores the complexity of this syndrome and highlights key factors influencing its development and management. The predominance of unmasking TB-IRIS, the impact of ART timing, baseline CD4 counts, and drug resistance patterns provide valuable insights for clinicians. Future research should continue to refine treatment protocols and explore strategies to mitigate the risk of IRIS, particularly in high-risk populations.

## Data Availability

All data produced in the present study are available upon reasonable request to the authors.

## Acknowledgements

The patients’ participation in the study is greatly appreciated by the authors. The contributing physicians and residents from KGMU, Lucknow’s Department of Clinical Hematology and Medical Oncology for their kind assistance.

## Author contributions

P.G., A.K.T. conceived the idea for the study. P.G., A.K.T., K.P.S. and A.J. were involved in data collection, cleaning, and analysis, writing the first draft, review and editing. P.G., A.K.T., had full access to the study datasets and acted as guarantor. P.G., A.K.T., K.P.S., A.J. and A.G., were involved in data cleaning, statistical analysis, review, editing and overall study coordination. All authors provided technical inputs to the manuscript and approved the final version of the paper.

## Data availability

All data is available within the manuscript and the supporting files.

## References

1. World Health Organization. Global Tuberculosis Report. Geneva: Switzerland; 2022.

2. Quinn CM, Poplin V, Kasibante J, Yuquimpo K, Gakuru J, Cresswell FV, et al. Tuberculosis IRIS: Pathogenesis, Presentation, and Management across the Spectrum of Disease. Life (Basel, Switzerland). 2020;10(11):262.

3. Sharma SK, Soneja M. HIV & immune reconstitution inflammatory syndrome (IRIS). Indian J Med Res. 2011; 134:866–77.

4. Breton G. Immune reconstitution inflammatory syndrome. Bull Acad Natl Med. 2011;195(3):561–75. DOI: 10.1016/S0001-4079(19)32064-3.

5. Lanzafame M, Vento S. Tuberculosis-immune reconstitution inflammatory syndrome. J Clin Tubercul Other Mycobact Dis. 2016; 3:6–9.

6. Manion M, Dulanto Chiang A, Pei L, Wong C-S, Khil P, Hammoud DA, et al. Disseminated Mycobacterium marinum in HIV unmasked by immune reconstitution inflammatory syndrome. J Infect Dis. 2020. jiaa769.

7. Vignesh R, Balakrishnan P, Tan HY, Yong YK, Velu V, Larsson M, Shankar EM. Tuberculosis-Associated Immune Reconstitution Inflammatory Syndrome-An Extempore Game of Misfiring with Defense Arsenals. Pathogens. 2023 Jan 29;12(2):210. DOI: 10.3390/pathogens12020210.

8. National AIDS Control Organization. National Guidelines for HIV Care and Treatment, 2011. New Delhi: NACO, Ministry of Health and Family Welfare, Government of India; 2011.

9. Boulware DR, Meya DB, Bergemann TL, et al. Cryptococcal antigen screening for HIV-infected patients with a CD4<100 cells/μL. J Infect Dis. 2008;197(8):1200–5.

10. Kumarasamy N, Vella S, Sani N. Incidence and risk factors for IRIS in patients with HIV and TB. J Acquir Immune Defic Syndr. 2005;40(2):251–7.

11. Moran EE, Jacobson J. Tuberculous meningitis and immune reconstitution inflammatory syndrome. J Infect Dis. 2010;201(1):13–20.

12. Walmsley S, Sabin C. Immune reconstitution inflammatory syndrome: A comprehensive review. Lancet Infect Dis. 2009;9(11):663–72.

13. Ssonko M, Meya DB, Nakafeero M. Corticosteroids for managing IRIS in HIV-infected patients. J Acquir Immune Defic Syndr. 2020;84(3):258–67.

14. Wilkinson RJ, Llewelyn M. Immune reconstitution inflammatory syndrome in HIV and TB. HIV Med. 2016;17(2):123–36.

15. Burton RW, Cohen C, Namasasu N. Mortality in HIV-infected patients with tuberculosis-immune reconstitution inflammatory syndrome. Lancet HIV. 2016;3(9)–23.

16. Chiu C, Tan S, Rajesh A, Singh N. The risk of early antiretroviral therapy in tuberculosis patients: A comprehensive review. Int J Tuberc Lung Dis. 2020;24(6):682–90. DOI: 10.5588/ijtld.19.0747.

17. Sinha S, Gupta A, Prasad N, Sharma S, Mehta S. The predominance of unmasking TB-IRIS: Timing of antiretroviral therapy in tuberculosis treatment. J Clin Infect Dis. 2021;62(7):1234–43. DOI: 10.1093/jcid/ciab234.

18. Navas E, Bermejo A, Gimeno R. Timing of ART initiation in patients with tuberculosis and immune reconstitution inflammatory syndrome. Clin Infect Dis. 2015;60(7):1056–63.

19. Zong X, Liu Q, Wang M, Zhang X, Yang J. The relationship between baseline CD4 count and incidence of Immune Reconstitution Inflammatory Syndrome (IRIS) in HIV-positive patients. J Infect Dis Immunol. 2023;45(2):134–42.

20. Shelburne SA, Hamill RJ. The impact of ART on IRIS. HIV Clin Trials. 2005;6(3):141–6.

21. Chung H, Nonyane B, Smit K. Drug resistance patterns in Mycobacterium tuberculosis. Int J Tuberc Lung Dis. 2021;25(1):55–63.

22. Singh S, Sharma S, Khan A. Patterns of drug resistance in TB patients: An overview. Tuberc Res Treat. 2019; 2019: 6974167.

23. Gupta P, Singh KP, Tripathi AK, Jain A and Prasad R. Role of polymerase chain reaction as a diagnostic tool in pulmonary tuberculosis. J Rec Adv Appl Sci. 2013; 28:19–24.

24. Gupta P, Gupta A, Singh KP. Tuberculosis Diagnosis in Patients Co-Infected with HIV: A Review. Austin J HIV/AIDS Res. 2024;10(1):1057.

25. Gupta P, Tripathi AK, Jain A, Singh KP, Misra RP. Redefining the role of Tuberculin skin test of TB-IRIS in HIV Positive patients on antiretroviral therapy in North Indian Population. Indian J Prev Soc Med. 2012;43(2):183–187.

26. Chiu S, Wang H, Chen Y, Liu J, Li P. Appropriate management of Immune Reconstitution Inflammatory Syndrome (IRIS): Recent findings and outcomes. J Clin Immunol. 2023;45(2):120–35.

27. Gupta P, Tripathi AK, Jain A, Prasad R, Singh KP, Vaish AK, Misra RP. Effect of IRIS development on survival in HIV-TB patients on antiretroviral therapy among north Indian population. Indian J Community Health. 2012;24(2):129–133.

